# Ambient nitrogen dioxide pollution and spread ability of COVID-19 in Chinese cities

**DOI:** 10.1101/2020.03.31.20048595

**Authors:** Ye Yao, Jinhua Pan, Zhixi Liu, Xia Meng, Weidong Wang, Haidong Kan, Weibing Wang

**Affiliations:** Fudan University, Shanghai China

## Abstract

The Coronavirus (COVID-19) epidemic, which was first reported in December 2019 in Wuhan, China, has caused 219,331 confirmed cases as of 20 March 2020, with 81,301 cases being reported in China. It has been declared a pandemic by the World Health Organization in 11 March 2020 (1). Although massive intervention measures have been implemented in China (e.g. shutting down cities, extending holidays and travel ban) and many other countries, the spread of the disease are unlikely to be stopped over the world shortly. It is becoming evident that environmental factors are associated with seasonality of respiratory-borne diseases’ epidemics (2). Previous studies have suggested that ambient nitrogen dioxide (NO_2_) exposure may play a role in the phenotypes of respiratory diseases, including, but not limited to, influenza, asthma and severe acute respiratory syndrome (SARS). NO_2_), for example, might increase the susceptibility of adults to virus infections (3). High exposure to NO_2_ before the start of a respiratory viral infection is associated with the severity of asthma exacerbation (4). This study aims to assess the associations of ambient NO_2_ levels with spread ability of COVID-19 across 63 Chinese cities, and provides information for the further prevention and control of COVID-19.

## Methods

We collected COVID-19 confirmed case information reported by the National Health Commission and the Provincial Health Commissions of China.

We calculated basic reproduction number (R_0_) for 63 cities with more than 50 cases as of February 10 (COVID-19 peak time in China, including 12 cities in Hubei and 51 cities outside Hubei). The R_0_ means the expected number of secondary cases produced by an initial infectious individual, in a completely susceptible population. The calculation process is completed by R software.

Hourly NO_2_ data were obtained from the National Urban Air Quality Publishing Platform (http://106.37.208.233:20035/), which is administered by China’s Ministry of Environmental Protection. Daily concentrations of NO_2_ were calculated as the average of at least 18 (75%) hourly concentrations for all state-controlled stations, then daily NO_2_ levels of the city was averaged from all valid stations within it. Other meteorological data including daily mean temperature and relative humidity were collected from the China Meteorological Data Sharing Service System.

We conducted a cross-sectional analysis to examine the spatial associations of NO_2_ with R_0_ of COVID-19, and a longitudinal analysis to examine the temporal associations (day-by-day) of NO_2_ with R_0_ in the cities in Hubei province since they had enough confirmed case number to acquire stable daily R_0_ and the other covariates including health policies were quite similar inside Hubei. We used multiple linear regression to assess the relationship between the spread ability of COVID-19 and NO_2_ pollution across the different cities.

## Results

Among 63 cities, the mean ± standard deviation and range were (27.9±8.3, 10.7-53.0) for NO_2_ and (1.4±0.3, 0.6-2.5) for R_0_. The top three cities (Wuhan, Huanggang and Yichang) with the highest R_0_ were all in Hubei Province.

The cross-sectional analysis shows that, after adjustment for temperature and humidity, the R_0_ was positively associated with NO_2_ in all cities (χ ^2^=10.18, *p*=0.037). In a following stratified analysis, a significant association was confirmed in the cities outside of Hubei (r=0.29, *p*=0.046), while it is not the case in the cities inside Hubei (r=0.51, *p*=0.130) (Figure 1). We did not find signification associations of temperature and relative humanity with R_0_ of COVID-19 (χ^2^=4.62, *p*=0.372 and χ ^2^=1.63, *p*=0.804).

**Figure 1.**
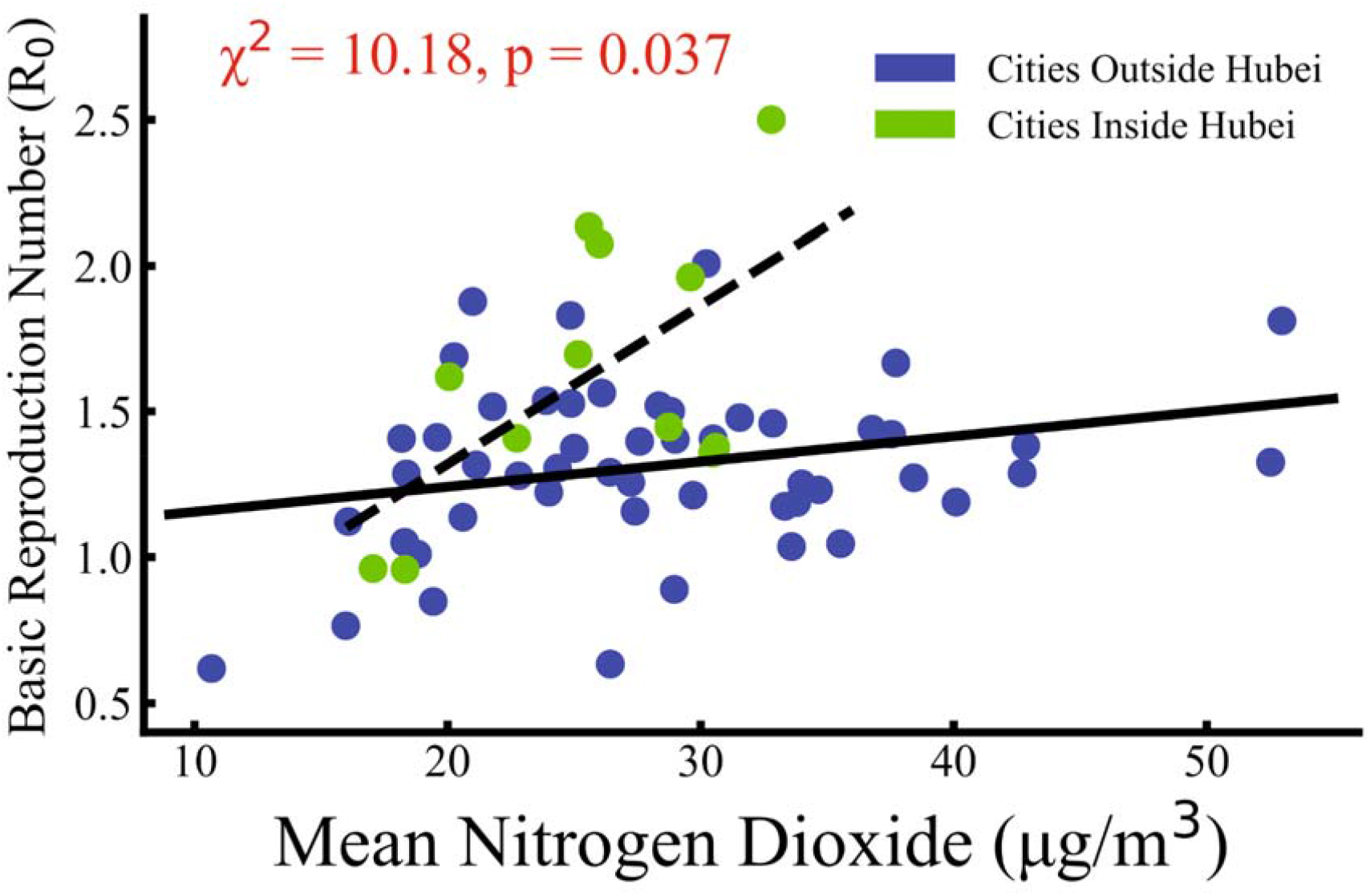
Nitrogen Dioxide and Spread Ability of COVID-19 in Spatial Scale. Basic Reproduction Number R_0_ was positively associated (Meta χ^2^=10.18, p=0.037) with NO_2_ in cities outside Hubei (blue points, 51 cities, r=0.29, p=0.046) and cities inside Hubei (green points, 12 cities, r=0.51, p=0.13). Temperature and humidity effects have been removed during statistical analysis.

In temporal scale, we calculated daily R_0_ of 11 cities in Hubei except Wuhan from January 27 to February 26 (there were few COVID-19 confirmed cases in these cities afterwards), and normalized them based on Wuhan’s daily R_0_ in order to avoid other covariates’ effects. We found that the 11 Hubei cities (except Xianning City) all held significant positive correlations between NO_2_ (with 12-day time lag) and R_0_ (r>0.51, *p*<0.005), suggesting a positive association between NO_2_ and COVID-19 spread ability in the temporal scale (Figure 2).

**Figure 2.**
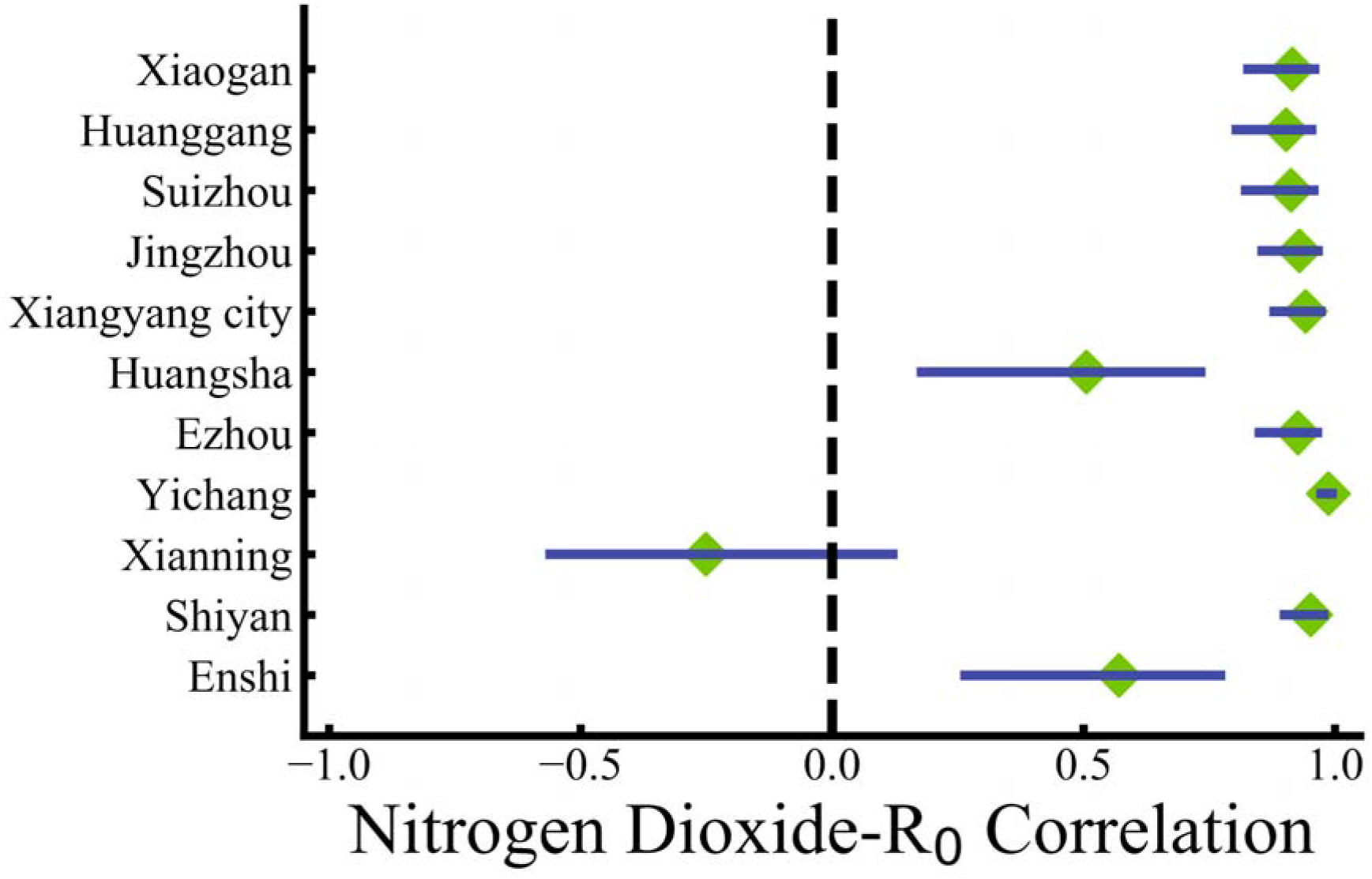
Nitrogen Dioxide and Spread Ability of COVID-19 in Temporal Scale in Hubei. Nitrogen Dioxide-R_0_ Temporal Correlation in 11 Hubei Cities. The 11 Hubei cities except Xianning all held significant positive correlations (r>0.51, p<0.005) between NO_2_ (with 12-day time lag) and daily R_0_ (normalized based on Wuhan’s daily R_0_).

## Discussion

Our study was designed to explore the association between the environment factors and the transmission of COVID-19. To our knowledge, this is the first study to investigate the ambient air pollution associated with the transmission of COVID-19. Our results reported the significant association between NO_2_ exposure and R_0_, suggesting that ambient NO_2_ may contribute to the spread ability of COVID-19. Previous studies have suggested that the increase spread ability from NO_2_ might not be caused by increased susceptibility of the epithelial cells to infection but may result from effects of NO_2_ on host defenses that prevent the spread of virus (5). Since NO_2_ is a traffic-related air pollutant, the association may also be explained by the relationship between virus spread and population movement. Clearly, further investigations are warranted to provide additional details and illustrate the mechanism.

Our study has limitations. Given the ecological nature of study, other city-level factors, such as implementation ability of COVID-19 control policy, urbanization rate, and availability of medical resources, may affect the transmissibility of COVID-19 and confound our findings. Future studies should develop individual based models with high spatial-temporal resolution to assess the correlation between air pollution and epidemiologic characteristics of COVID-19.

## Data Availability

COVID-19 confirmed cases and deaths information be obtained from the following link(The first and second link);Daily PM2.5 and PM10 data were obtained from the National Urban Air Quality Publishing Platform(The third link);

http://www.nhc.gov.cn/xcs/xxgzbd/gzbd_index.shtml

http://wjw.hubei.gov.cn/bmdt/ztzl/fkxxgzbdgrfyyq/

http://106.37.208.233:20035/

## Author contributions

Ye Yao, Weibing Wang, and Haidong Kan designed the study. Jinhua Pan, Zhixi Liu, Ye Yao, and Weibing Wang collected COVID-19 incidence data and gained insight into the biology and natural history of the virus. Jinhua Pan, Zhixi Liu., Ye Yao and Weibing Wang developed the model and obtained the related parameters. Weidong Wang and Haidong Kan collected meteorological factors. Ye Yao, Jinhua Pan, Zhixi Liu, and Xia Meng drafted the manuscript. Haidong Kan and Weibing Wang revised the manuscript. All authors critically reviewed and approved the final version of the manuscript.

## Competing interests

The authors declare no competing interests.

## Acknowledgements

This study was sponsored by the Bill & Melinda Gates Foundation (Grant No. OPP1216424) and Fudan University Research Project on COVID-19 Emergency (Grant No. IDF201007).

## Supplementary Material

### Data collection

We collected COVID-19 confirmed case information in China reported by the National Health Commission and the Provincial Health Commissions of China. We calculated basic reproduction number (R_0_) for 63 cities with more than 50 cases as of February 10 (including 12 cities in Hubei and 51 cities outside Hubei).

### Data analysis

We conducted a cross-sectional analysis to examine the spatial associations of NO_2_ with R_0_ of COVID-19, and examined the temporal day-by-day associations of NO_2_ with R_0_ in cities of Hubei province since they had enough confirmed case number to acquire stable daily R_0_ and the other covariates including health policies were quite similar inside Hubei. We used multiple linear regression to assess the relationship between the spread ability of COVID-19 and nitrogen dioxide pollution across different cities. The basic reproduction number, denoted R_0_, means the expected number of secondary cases produced by an initial infectious individual, in a completely susceptible population. if R_0_ < 1, then the disease free equilibrium is locally asymptotically stable; whereas if R_0_ > 1, then it is unstable. Thus, R_0_ is a threshold parameter. The calculation process is completed by R software.

### Spatial distribution of NO_2_

**Supplementary Figure 1.**
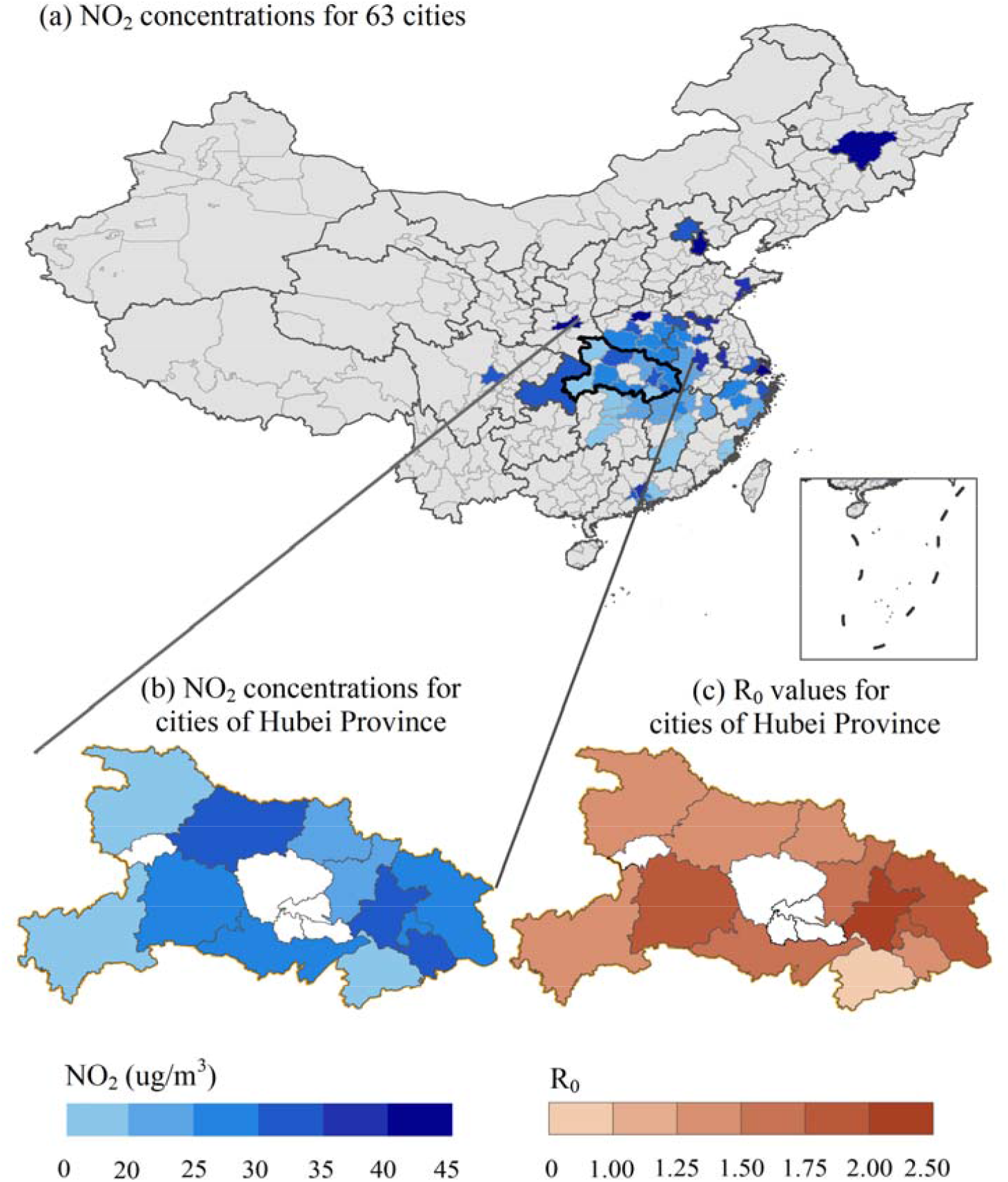
The spatial Distribution of Average of Nitrogen Dioxide Concentration and Spread Ability of COVID-19. The China map shows the spatial distribution of the average nitrogen dioxide concentration from January 1, 2020, to February 8, 2020, in 63 Chinese cities. And zoom up the “Hubei province” part to compare the trend of the average nitrogen dioxide concentration (gradient blue map, bottom left) with the spatial trend of the basic reproduction number R_0_ (gradient brown map, bottom right) in Hubei province.

## Notes

### Competing Interest Statement

The authors have declared no competing interest.

